# Predictive value of EEG/ECG Biomarkers for Treatment Response in Depression

**DOI:** 10.64898/2026.03.25.26349315

**Authors:** Barbora Provaznikova, Mateo de Bardeci, Enzo Altamiranda, Cheng-Teng Ip, Anna Monn, Samantha Weber, Johannes Jungwirth, Judith Rohde, Susanne Prinz, Golo Kronenberg, Annette Brühl, Tobias Bracht, Sebastian Olbrich

**Affiliations:** Department of Psychiatry, Psychotherapy and Psychosomatics, University Hospital of Psychiatry, Zürich, Lenggstrasse 31, 8032 Zürich, Switzerland; DeepPsy AG, Forchstrasse 154, 8125 Zollikerberg, Switzerland; Centre for Cognitive and Brain Sciences, University of Macau, Taipa, Macau SAR, China; Department of Electrical and Computer Engineering, Faculty of Science and Technology, University of Macau, Taipa, Macau SAR, China; University Psychiatric Clinics Basel, Clinic for Adults, University of Basel, Basel, Switzerland; Competence Center for Interventional Psychiatry and Augmented Psychotherapy, University Hospital of Psychiatry and Psychotherapy, University of Bern, Bern, Switzerland

**Keywords:** Major depressive disorder, EEG, ECG, Biomarkers, Personalized Psychiatry, Treatment response prediction

## Abstract

**Objective:** Major depressive episodes frequently show limited response to first-line treatments, motivating the search for objective biomarkers. EEG/ECG-based support tools aggregating electrophysiological predictors may guide treatment selection. We examined whether antidepressant treatments concordant with an EEG/ECG-biomarker report were associated with higher response rates.

**Methods:** We retrospectively analyzed adults with ICD-10 depressive disorder or bipolar depression treated with electroconvulsive therapy (ECT), repetitive transcranial magnetic stimulation (rTMS), (es)ketamine, or selective serotonin reuptake inhibitors (SSRIs) between 2022 and 2024. Resting-state EEG with simultaneous ECG generated individualized biomarker reports with modality-specific response likelihoods. Treatment chosen by clinical teams was classified as concordant or non-concordant; response was derived from routinely collected clinical scales.

**Results:** Among 153 patients (ECT n=53, rTMS n=48, (es)ketamine n=36, SSRIs n=16), response rates were higher for concordant vs non-concordant treatments: ECT 70% vs 50%, rTMS 30% vs 13%, (es)ketamine 31% vs 10%, and SSRIs 100% vs 11%. Overall, 46% (42/92) of concordant vs. 26% (14/54) of non-concordant patients responded (absolute difference +20 percentage points; relative increase ≈77%; number needed to treat ≈5).

**Conclusion:** Concordance with EEG/ECG biomarkers correlated with higher treatment response, warranting confirmation in prospective trials.

**Significance:** EEG/ECG-based decision support may enhance antidepressant treatment response in everyday clinical practice.

## Introduction

Major depressive disorder (MDD) is a leading cause of disability worldwide, currently affecting approximately 280 million individuals (Friedrich, 2017) with associated social and economic burden (Wang et al., 2006). Despite guideline-recommended evidence-based pharmacological, psychotherapeutic, and interventional treatments, remission with a first adequate antidepressant trial typically reaches about 30-40%, and even overall response rates remain below 50%, so that a substantial proportion of patients remain symptomatic after several weeks of treatment (Belanger et al., 2023; Rush et al., 2006). In everyday practice, treatment often involves successive medication switches, dose titrations, and augmentation strategies that depend on clinician experience and patient preference, leading to lengthy and expensive care (DGPPN et al., 2017).

In response, psychiatric research has increasingly aimed for precision and stratified medicine, seeking to use objective markers to individualize treatment selection rather than applying “one size fits all” algorithms (Arns et al., 2022). Clinical and questionnaire-based prediction models that combine symptom scales, sociodemographic factors, and routine clinical variables can improve the prediction of non-response or treatment resistance to standard antidepressants, but their individual level discriminative performance remains moderate (Chekroud et al., 2017; Perlis, 2013). These findings suggest that clinical and questionnaire-based models may not be sufficient to guide individual treatment decisions. In parallel, algorithm-guided decision support approaches based on structured clinical and trial data have been shown to increase remission rates, shorten time to remission, and improve cost-effectiveness compared with usual care in major depression (Adli et al., 2017; Bauer et al., 2019; Ricken et al., 2011).

Resting-state-electroencephalography (EEG) has been investigated as a potential tool for treatment prediction that can be derived from standard hospital equipment. Meta-analyses emphasize substantial heterogeneity and conclude that EEG markers are not yet sufficiently reliable for routine clinical prediction in depression. Nevertheless, several multicenter studies report that features such as alpha asymmetry (reflecting the balance of left-right frontal activity) and vigilance regulation are associated with response to selective serotonin reuptake inhibitors (SSRIs), serotonin–norepinephrine reuptake inhibitors (SNRIs), and repetitive transcranial magnetic stimulation (rTMS), with replication across independent cohorts (Arns et al., 2016; Olbrich and Arns, 2013; Widge et al., 2019). Most recently, a large deep learning study using multicenter resting-state EEG data demonstrated-that EEG-based models can diagnose MDD and predict SSRI treatment response with accuracies around 70–80% (Arns et al., 2016; Olbrich et al., 2016a). In the iSPOT D trial, alpha asymmetry was validated as a sex-specific predictor of differential antidepressant response and combined central nervous system and autonomic nervous system measures further increased the probability of response and remission (Meyer et al., 2021a; Olbrich et al., 2016a). Autonomic indices derived from electrocardiography (ECG), such as heart rate variability or heart-rate dynamics at rest, have likewise been linked to response to (es)ketamine and conventional antidepressants and may enhance predictive accuracy when combined with EEG features (Meyer et al., 2021a; Olbrich et al., 2016a). In addition to these individual findings, growing evidence has characterized EEG and ECG markers as predictors of antidepressant treatment response across different drug classes and neuromodulation protocols, including large, randomized trials and independent replication studies. This literature, encompassing alpha peak frequency, alpha asymmetry, vigilance regulation, heart rate and heart_rate variability, and combined CNS-ANS arousal measures, forms the empirical basis for EEG/ECG based biomarker signatures that can be integrated into clinical decision support tools, such as a tool called DeepPsy. (Arns et al., 2018, 2018, 2017; Badrakalimuthu et al., 2011; Corlier et al., 2019; Dohrmann et al., 2017; Ferreira-Garcia et al., 2021; Geissler et al., 2014; Hegerl et al., 2008; Hegerl and Hensch, 2014; C. Ip et al., 2024, 2021; C.-T. Ip et al., 2021; Meyer et al., 2021a; Olbrich et al., 2022, 2016b, 2012a, 2012b; Roelofs et al., 2021; Rüesch et al., 2023; Schumann and Bär, 2022; Stoppe et al., 2019; Ulke et al., 2019; van der Vinne et al., 2021, 2019; Voetterl et al., 2023).

In our clinical setting, EEG and ECG recordings are processed using the DeepPsy system, which combines predefined electrophysiological markers with published treatment outcome associations to yield modality specific likelihoods of response for guideline recommended interventions in depression (DeepPsy AG, 2025), consistent with current national and international clinical guidelines and regulatory frameworks (Bundesärztekammer (BÄK), Kassenärztliche Bundesvereinigung (KBV), et al., 2022; DGPPN et al., 2017; Hättenschwiler et al., 2016).

The present work aimed to evaluate an EEG/ECG based biomarker report as an aid for horizontal treatment decisions. The report is not intended to determine which line of treatment (e.g. antidepressants, (es)ketamine, rTMS or electroconvulsive therapy (ECT)) is indicated (vertical treatment decision), but to provide additional information for choosing between guideline recommended options within a given line of care, such as which antidepressant, which rTMS protocol, or whether ECT or (es)ketamine may have a higher likelihood of response. Specifically, we examined in routine clinical practice whether treatment decisions classified as concordant with the biomarker report were associated with higher response rates than non-concordant decisions across four modalities (ECT, rTMS, (es)ketamine, SSRIs) in a pooled analysis. We hypothesized that, within each modality, patients receiving concordant treatment would show higher response rates than those receiving non-concordant treatment, and that, across all modalities, concordant treatment would overall be associated with higher response rates. To quantify the magnitude and potential clinical relevance of these effects, we compared response proportions and derived relative improvements and numbers needed to treat (NNT) across and within modalities.

## Methods

### Setting and design

This retrospective analysis was conducted at the University Hospital of Psychiatry in Zürich, Switzerland. Adult patients (≥18 years) with a current MDD or depressive episode within a bipolar disorder (BD) according to ICD-10 criteria were treated in routine in and outpatient care with one of four guideline-based interventions for depression: SSRIs, (es)ketamine, rTMS, or ECT. In accordance with standard clinical practice, patients receiving neuromodulation or (es)ketamine interventions had undergone at least two prior antidepressant and/or augmentation trials without sufficient response. Depressive episode severity ranged from mild to severe. Diagnostic assessment and treatment indication were established by the treating psychiatrist and confirmed by at least one supervising psychiatrist. Patients were eligible for the present analysis if they received exactly one of these four modalities, had baseline EEG/ECG recordings, and had complete pre- and posttreatment outcome ratings for the respective modality. The final sample comprised 153 patients: 53 received ECT, 48 rTMS, 36 (es)ketamine, and 16 SSRIs.

### Concordant versus non-concordant treatment

For each treatment, the EEG/ECG report (DeepPsy) provides a treatment specific likelihood of response. Treatment was classified as report-concordant when the administered modality (and, for rTMS, the delivered protocol) corresponded to a DeepPsy recommendation indicating increased likelihood of response. Treatment was classified as non-concordant when a modality was chosen while the report did not indicate increased likelihood of response for that modality. For all four modalities, response rates were compared between report-concordant and non-concordant groups. In routine care, deviations from DeepPsy recommendations could occur for several reasons, including lack of use of the report in individual cases, patient preferences regarding specific modalities, or clinical considerations such as prior nonresponse or contraindications that led clinicians to select an alternative treatment. Accordingly, non-concordant decisions do not necessarily reflect disagreement with the report but rather the integration of biomarker information into broader clinical decision making.

### Electroconvulsive therapy (ECT)

Patients in the ECT group received a standard course of 12 sessions according to established hospital protocols. Treatments were administered with bifrontal electrode placement under general anesthesia using standard parameters that indicated a qualitatively sufficient seizure (de Arriba-Arnau et al., 2021; Exner et al., 2023; Kranaster et al., 2018). Symptom severity was assessed before and after treatment using the Clinical Global Impression scale (CGI) (Busner and Targum, 2007). All CGI ratings were conducted by 2 clinically experienced physicians. When disagreement occurred between the 2 raters, all available data were used to make a final agreement rating. The CGI subscale efficacy index (CGI-E) served as primary outcome to define response (R) and nonresponse (NR). It compares the patient’s initial state to a ratio of current therapeutic benefit and severity of side effects. Response thereby was defined by a CGI-E score of 1 or 2 (1 = vast improvement, complete or nearly complete remission of all symptoms, 2 = decided improvement, partial remission of symptoms). Nonresponse was marked by CGI-E scores of 3 or 4 (3 = slight improvement, which does not alter status of care of patient, 4 = unchanged or worse). For evaluating the effectiveness of the DeepPsy recommendation for ECT treatment, clinical response was compared for the group that received ECT following the DeepPsy report and the group of patients that received ECT although DeepPsy recommendation did not state an increased likelihood of response. In total, 50 patients were included into the cohort, from whom CGI, EEG- and ECG baseline data were available.

### Repetitive Transcranial magnetic stimulation (rTMS)

To evaluate the DeepPsy recommendation for rTMS protocol selection, we included patients who received 1Hz stimulation over the right dorsolateral prefrontal cortex (rDLPFC). We classified them as report-concordant when 1Hz was recommended by DeepPsy, and as non-concordant when the 1Hz protocol was chosen even though another protocol was recommended. Patients in the rTMS group received 20 sessions of 1Hz rTMS over the right dorsolateral prefrontal cortex (rDLPFC), with 600–1200 pulses per session (Lefaucheur et al., 2020) and 3-5 sessions per week. Depressive symptoms were measured with the Beck Depression Inventory II (BDI II) (Beck et al., 1961). Response was defined as a ≥50% reduction in BDI II score from baseline. 48 patients with complete BDI II, EEG, and ECG data were included.

### (Es)Ketamine

Patients received intranasal esketamine up to 84 mg per session or racemic ketamine up to 100 mg per session, for a total of 12 sessions over 8 weeks (two sessions per week for the first 8 sessions and one session per week for the final 4 sessions). Symptom severity was assessed with the BDI II (Beck et al., 1961) and response was defined as a ≥50% reduction from baseline. In total, 36 patients with complete BDI II, EEG, and ECG baseline data were included.

### Selective Serotonin Reuptake Inhibitors

Data was received from 16 patients who were treated with escitalopram (10–20 mg/day) or sertraline (50–200 mg/day) as first-line pharmacotherapy over a 6 week period. Depressive symptoms were assessed using the Hamilton Depression Rating Scale (HAMD) (Hamilton, 1960), and response was defined as a ≥50% reduction in total HAMD score from baseline. Therapeutic drug monitoring was not performed.

### EEG/ECG acquisition

Before treatment initiation, all participants underwent resting!ZIstate EEG with simultaneous ECG recordings in a standard clinical setting. EEG was recorded with a 22-channel montage positioned according to the international 10-20 system, including the following scalp electrodes: Fp1, Fp2, F3, F4, C3, C4, P3, P4, O1, O2, F7, F8, T3, T4, T5, T6, Fz, Cz, Pz, Oz and a reference electrode. Impedances were kept below 5Ω. Signals were recorded using a Neurofax 1200 amplifier (Nihon Kohden), together with a bipolar ECG montage attached to the left and right wrists. For this analysis, 2–6 minutes of artefact!ZIfree, eyes!ZIclosed resting!ZIstate EEG were used, sampled at 250–1000 Hz. Preprocessing was performed using DeepPsy software (version v0.3.167MD; DeepPsy Biomarkers, v1.02, 2024) and comprised segmentation, manual artefact rejection, independent component analysis (ICA), cascade filtering (high!ZIpass 0.01 Hz, low!ZIpass 70 Hz, 50 Hz notch), and re!ZIreferencing to the grand average. For the ICA, up to four independent components per recording, typically corresponding to ocular or movement-related artefacts, were removed based on visual inspection by trained raters using standardized criteria. The processed EEG and ECG features were then entered into the DeepPsy algorithm to generate individual biomarker reports with treatment!ZIspecific response likelihoods for ECT, rTMS, (es)ketamine, and SSRIs.

### DeepPsy biomarker report

The DeepPsy report is an EEG/ECG-biomarker report derived from a systematic review of the predictive electrophysiological literature published over the last 12 years. It integrates predefined EEG and ECG biomarkers with published treatment outcome associations to support treatment selection in psychiatric care. The report presents quantitative biomarker values together with literature derived associations to treatment response. Note that the report does not provide diagnostic classifications.

Treatment specific rules are derived from peer reviewed studies linking EEG or ECG markers to differential treatment response and are annotated with an evidence level. In a rule based integration step, each biomarker contributes a positive, negative, or neutral weight to each evaluated treatment modality (SSRIs or SNRIs, rTMS, (es)ketamine and ECT). Weights are combined across biomarkers, taking into account the reported level of evidence for each association, to generate modality specific likelihood of response scores.

Relevant EEG biomarkers include vigilance regulation and vigilance level measures, alpha peak frequency, and frontal alpha asymmetry. These markers have been reported to show differential associations with treatment response across treatment modalities. ECG derived biomarkers include resting heart rate, heart rate dynamics over time, and heart rate variability measures reflecting autonomic nervous system regulation, including high frequency power, low frequency power, and the LF HF ratio, which have been associated with treatment outcomes particularly in pharmacological and ketamine-based interventions (Ferreira-Garcia et al., 2021; Meyer et al., 2021a; Olbrich et al., 2022, 2016b, 2016b). In a pre-defined, rule-based integration step, each biomarker contributes a positive, negative, or neutral weight to each evaluated treatment modality. Weights are combined across biomarkers while accounting for the reported level of evidence for each association to generate modality specific likelihood of response scores.

The final DeepPsy report presents quantitative EEG and ECG biomarker values with normative reference ranges, narrative interpretations linked to published evidence, and aggregated modality specific likelihood of response categories (for example increased likelihood of response, normal efficacy, or reduced likelihood). If applicable, these outputs can be used as decision support information alongside clinical judgement in routine care settings.

### Statistical analysis

Response rates between treatment decisions that were concordant versus non-concordant with the DeepPsy report were compared using two complementary statistical approaches. Fisher’s exact test was applied for treatment groups with small or unbalanced sample sizes, as it provides exact p-values without relying on large-sample assumptions. For larger groups, including the combined dataset, we used a two-proportion z-test to provide a robust approximation of differences in responder proportions. All tests were performed two-sided and one-sided to evaluate both strict significance and the directional hypothesis that following the report improves treatment response. It is argued, that the one-sided test is used to judge on the effectiveness of the DeepPsy report in increasing the treatment response rate, since the assumption is built in literature in the efficacy of all used markers in the DeepPsy (Arns et al., 2023, 2016; C.-T. Ip et al., 2024; Meyer et al., 2021b; Monn et al., 2025; Olbrich et al., 2016a; Voetterl et al., 2024).

### Missing data

Cases with missing baseline EEG/ECG data or missing primary outcome ratings were excluded from the respective analyses; final sample sizes for each treatment arm are reported in the Results and in Tables 2 (response counts) and 3 (statistical tests).

### Ethics

De_identified, routinely collected clinical parameters were extracted from patient health records, including age, sex, diagnoses, concurrent medication, psychometric assessment scores, EEG and ECG. Patients were informed in advance about the de_identification process and the intended use of their data for research, and clinicians explained the option to provide consent for secondary use of health_related data in detail. Only patients who provided written informed consent for the use of their anonymized clinical and biomarker data in research were included. All procedures complied with the Declaration of Helsinki and applicable national data protection regulations. An ethical vote was obtained by the Swiss Association of Research Ethics Committees (Nr. 2023-00494), and ethical approval was granted (Nr. 2025-00559).

## Results

### Sociodemographic data

Sociodemographic characteristics of the total cohort and treatment subgroups are summarized in Table 1. Mean age ranged from 41.7 years (SD 11.5) in the SSRI group to 51.2 years (SD 12.1) in the ECT group. The proportion of female patients ranged from 52.1% in the rTMS group to 66.0% in the ECT group, with intermediate values of 62.5% in the SSRI group and 63.9% in the (es)ketamine group.

**Table 1:**
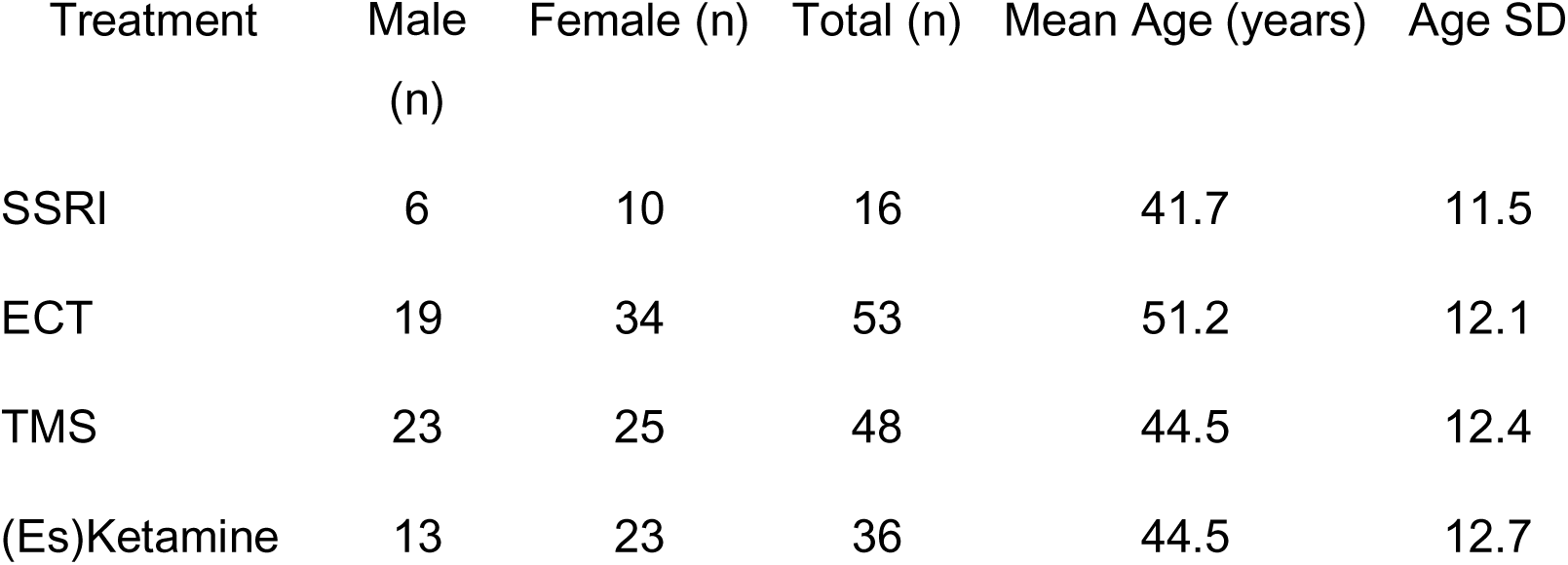
Demographic characteristics (age and gender) by treatment group.

| Treatment | Male<br>(n) | Female (n) | Total (n) | Mean Age (years) | Age SD |
| --- | --- | --- | --- | --- | --- |
| SSRI | 6 | 10 | 16 | 41.7 | 11.5 |
| ECT | 19 | 34 | 53 | 51.2 | 12.1 |
| TMS | 23 | 25 | 48 | 44.5 | 12.4 |
| (Es)Ketamine | 13 | 23 | 36 | 44.5 | 12.7 |

**Table 2:**
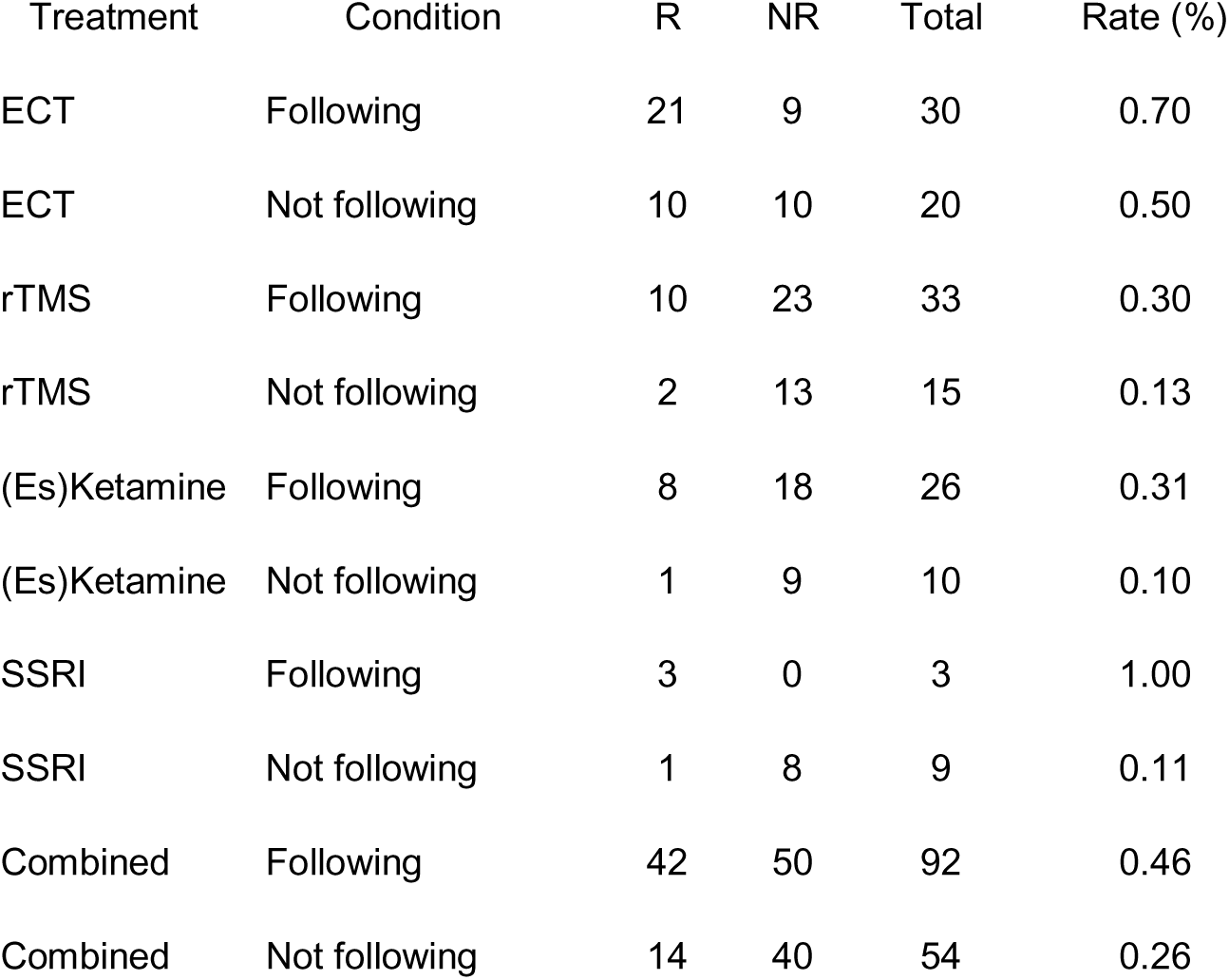
Numbers of responders and nonresponders for each treatment modality, stratified by report-concordant versus report-non-concordant use of the DeepPsy recommendations.

| Treatment | Condition | R | NR | Total | Rate (%) |
| --- | --- | --- | --- | --- | --- |
| ECT | Following | 21 | 9 | 30 | 0.70 |
| ECT | Not following | 10 | 10 | 20 | 0.50 |
| rTMS | Following | 10 | 23 | 33 | 0.30 |
| rTMS | Not following | 2 | 13 | 15 | 0.13 |
| (Es)Ketamine | Following | 8 | 18 | 26 | 0.31 |
| (Es)Ketamine | Not following | 1 | 9 | 10 | 0.10 |
| SSRI | Following | 3 | 0 | 3 | 1.00 |
| SSRI | Not following | 1 | 8 | 9 | 0.11 |
| Combined | Following | 42 | 50 | 92 | 0.46 |
| Combined | Not following | 14 | 40 | 54 | 0.26 |

### Treatment response by report concordance

In total, 153 patients were included (ECT n=53, rTMS n=48, (es)ketamine n=36, SSRIs n=16). Across all treatment modalities, treatments classified as concordant showed higher response rates than non-concordant treatments (Figure 1). Symptom ratings were obtained before treatment initiation and after completion of the treatment course for each modality (e.g. before the first ECT session and after 10-12 ECT sessions, before starting SSRI treatment and after 6 weeks, before the first rTMS session and after 20 sessions, and before the first (es)ketamine administration and after the 8-week treatment protocol).

**Figure 1:**
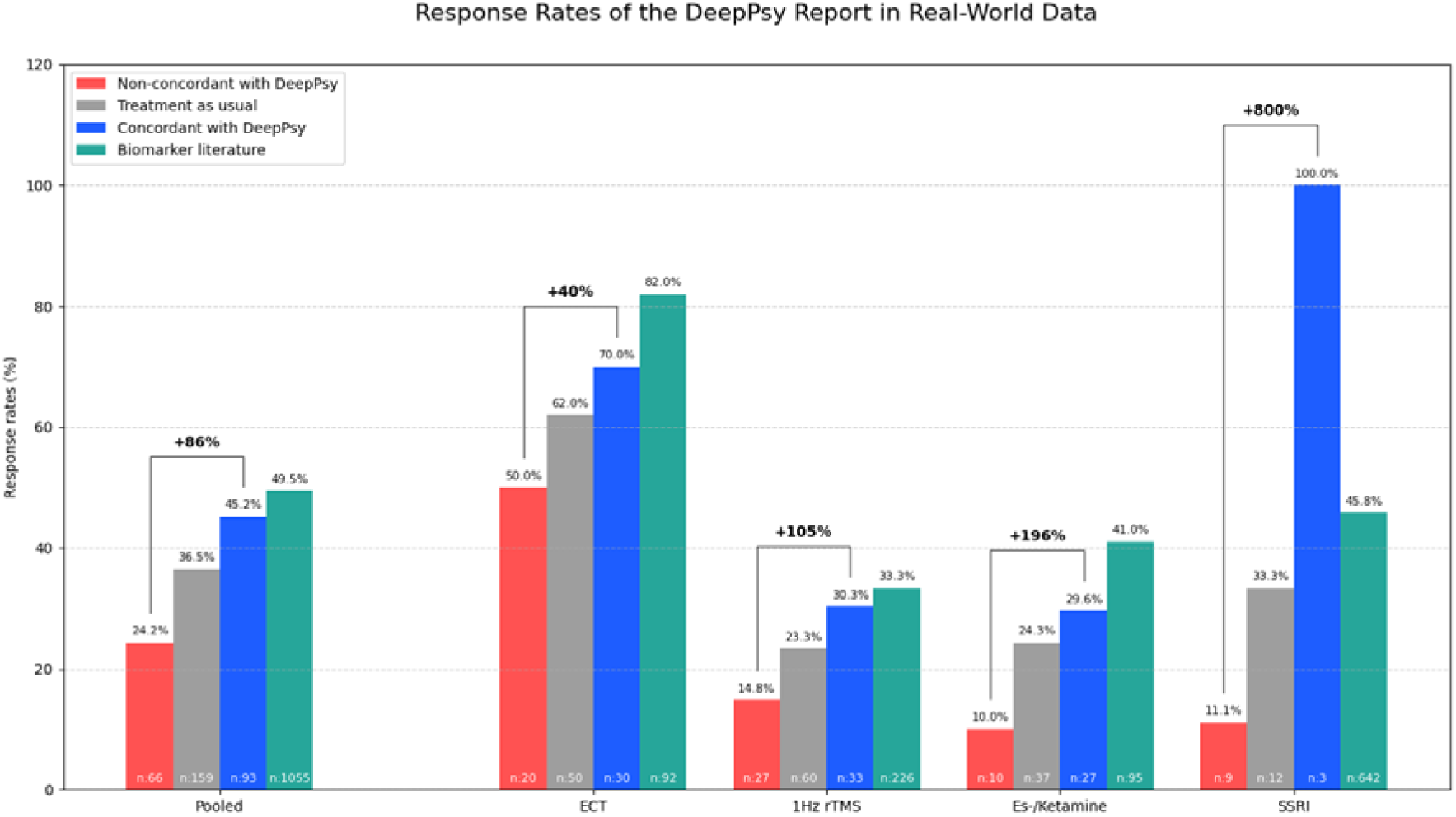
Treatment response rates to ECT, rTMS, (es)ketamine, and SSRI therapy categorized by DeepPsy non-concordant and DeepPsy concordant and biomarker literature. “Treatment as usual” (TAU) corresponds to the overall response rate observed in routine care for each modality, and “biomarker literature” indicates the expcted response range reported in prior EEG/ECG-biomarker studies for that modality.

In the ECT subgroup, response rates were 70% (21/30) with report-concordant treatment versus 50% (10/20) with non-concordant treatment, corresponding to an absolute risk difference of 20 percentage points (NNT≈5, one sided Fisher p=0.13; ztest p=0.08; OR 2.33, 95% CI 0.72–7.55; Table 3). For rTMS, response rates were 30% (10/33) versus 13% (2/15) (+17 percentage points; NNT≈6; Fisher p=0.19; ztest p=0.10; OR 2.83, 95% CI 0.54–14.92; Table 3). In the (es)ketamine group, response was 31% (8/26) for report-concordant treatment compared with 10% (1/10) for non-concordant treatment (+21 percentage points; NNT≈5; one sided z test p=0.10; OR 4.00, 95% CI 0.43–37.11; Table 3). The SSRI subgroup showed a large numerical difference, with response rates of 100% (3/3) versus 11% (1/9) (+89 percentage points; NNT≈1; Fisher p=0.02; z test p<0.01; OR 39.67, 95% CI 1.28–1229.87; Table 3), but this estimate is based on very small numbers and yields a wide confidence interval, so it should be interpreted with particular caution. When pooling all four modalities into a combined 2×2 comparison, report-concordant treatment was associated with 46% (42/92) responders compared with 26% (14/54) in the non-concordant group (+20 percentage points; relative increase ≈77%; NNT≈5; Fisher p=0.01–0.02; z test p=0.01–0.02; OR 2.40, 95% CI 1.15–5.00; Table 3).

**Table 3:** Statistical test results comparing antidepressant treatment response between report-concordant vs.report-non-concordant treatment. Subgroup analyses across individual treatment modalities showed higher responder proportions in the concordant groups, with statistically significant differences for SSRI treatment and for all treatments combined, and one-sided trends for ECT, rTMS, and (es)ketamine. *For the SSRI subgroup, the OR and confidence interval were computed using a Haldane–Anscombe correction (+0.5) due to the zero cell..

| Treatment | Rate followed | Rate not followed | $\Delta$ Rate | Odds ratio (OR) | 95% CI lower - 95% CI upper | Fisher p (1-sided) | Fisher p (2-sided) | Z-test p (1-sided) | Z-test p (2-sided) |
| --- | --- | --- | --- | --- | --- | --- | --- | --- | --- |
| ECT | 0.70 | 0.50 | 0.20 | 2.33 | 0.72-7.55 | 0.13 | 0.23 | 0.08 | 0.15 |
| TMS | 0.30 | 0.13 | 0.17 | 2.83 | 0.54-14.92 | 0.19 | 0.29 | 0.10 | 0.21 |
| (Es)Ketamine | 0.31 | 0.10 | 0.21 | 4.00 | 0.43-37.11 | 0.20 | 0.39 | 0.10 | 0.20 |
| SSRI | 1.00 | 0.11 | 0.89 | 39.67 | 1.28-1229.87 | 0.02 | 0.02 | <0.01 | <0.01 |
| Combined | 0.46 | 0.26 | 0.20 | 2.4 | 1.15-5.00 | 0.01 | 0.02 | 0.01 | 0.02 |

To interpret these effects in terms of absolute risk reduction and number needed to treat (NNT), we calculated NNTs from the observed risk differences. In the ECT subgroup, the response rate was 50% in the non-concordant group and 70% in the concordant group (+20 percentage points), corresponding to a relative increase of 40% and an NNT of 5 (1/0.20). For rTMS, response was 30% when concordant with the recommendations vs. 13% when non-concordant (+17 points; relative increase ≈130%; NNT≈6). In the (es)ketamine group, response in the concordant group was 31% compared to 10% in the non-concordant group (+21 points; relative increase ≈210%; NNT≈5). In the SSRI subgroup, response was 100% when concordant with the report vs. 11% when non-concordant (+89 points; NNT≈1), again with very small-numbers and wide uncertainty. Across all treatments combined, response increased from 26% in the non--concordant group to 46% in the concordant group (+20 points; relative increase ≈77%; NNT≈5). These NNTs in the range of approximately 5 may indicate clinically relevant effects, but they should be considered exploratory and interpreted in the light of the partly retrospective, non-randomized design and the small subgroup sizes, especially for SSRIs.

## Discussion

These findings suggest that treatments classified as concordant with the EEG/ECG-biomarker report were associated with higher response probabilities across all evaluated modalities in routine clinical practice. Absolute response improvements for ECT, rTMS and (es)ketamine ranged from +17 to +21 percentage points, corresponding to relative increases in response likelihood from approximately 40% (ECT) up to more than 200% ((es)ketamine) and yielding NNTs in the range of 5–6, which are comparable to, or better than, many established psychiatric interventions (Lefaucheur et al., 2020; Rush et al., 2006). The SSRI subgroup showed the numerically largest difference (+89 percentage points), although this estimate is based on small numbers and wide confidence intervals; nonetheless, the direction of effect was consistent across all treatments, and the combined analysis (46% vs 26% response) indicates an overall ∼20point absolute improvement and ∼77% relative increase in response associated with concordant treatment.

Importantly, these four evaluated modalities, ECT, rTMS, (es)ketamine, and SSRIs, cover a substantial part of the psychopharmacological and interventional spectrum of guideline-recommended treatments for depressive episodes that are approved in Switzerland and the European Union, which underscores the clinical context of these observations (DGPPN et al., 2017; Hättenschwiler et al., 2016; NICE guideline, 2022). To date, relatively few studies have evaluated EEG/ECG-based biomarker approaches for depression in real-world conditions; most work comes from controlled research settings or focuses on single biomarkers rather than integrated decision support tools (Arns, 2021; Widge et al., 2019). One of the few prospective applications is a feasibility study that used EEG features to guide choice among antidepressants, suggesting that biomarker-informed pharmacotherapy can be implemented in routine care (van der Vinne et al., 2021). Within this context, the present evaluation provides preliminary evidence that an EEG/ECG-based biomarker report may help to identify treatment options that are associated with higher response rates, while recognizing that causal effects cannot be inferred.

Previous work has shown that clinical and questionnaire-based prediction models derived from large antidepressant trials achieve moderate discrimination, and that individual EEG or ECG markers, while associated with treatment response, have not yet produced sufficiently robust single biomarker predictors for routine care (Chekroud et al., 2017; Perlis, 2013; Rush et al., 2006). Against this background, the present findings suggest that bundling multiple EEG/ECG features into an integrated decision-support report may yield clinically relevant effect sizes, but prospective validation will be required before firm conclusions can be drawn.

Several limitations should be considered when interpreting these findings. The retrospective, observational design means that the concordance of treatment with its recommendations were not experimentally controlled; reports were generated in routine care, and clinicians decided whether to consult and follow them, which introduces potential selection bias and confounding by indication that cannot be fully addressed. Unmeasured clinical factors may have influenced both the likelihood of obtaining a report and the decision to implement or override its suggestions, so that the observed associations cannot be interpreted as causal effects of biomarker guidance. Subgroup sample sizes, especially in the SSRI arm, were modest, resulting in wide confidence intervals and limiting formal statistical significance largely to the pooled analysis and the SSRI subgroup, with only suggestive trends for ECT, rTMS and (es)ketamine.

Further limitations concern the heterogeneity of outcome measures. In this naturalistic, retrospective sample, treatment response was assessed using different routinely employed instruments (CGI, BDI II, HAMD) rather than a single harmonized primary endpoint, reflecting ‘messy’ real-world documentation rather than a prespecified trial protocol. While each of these scales is widely used and has established validity for measuring depressive symptom change, their differing formats (clinician-rated vs self-report) and score ranges introduce additional measurement variability and complicate strict comparability across modalities. Notably, the CGI is a global illness severity and improvement rating rather than a depression specific symptom scale, which further contributes to heterogeneity in outcome assessment. As a result, the combined responder analyses should be interpreted cautiously, as they approximate a global clinical improvement construct rather than a uniform psychometric criterion of response. This heterogeneity is typical for observational data extracted from routine care and increases ecological validity, but it necessarily reduces the degree of standardization and statistical control compared with prospective trials.

Another constraint is that the algorithms underlying the biomarker report are proprietary: although they are grounded in an extensive published EEG/ECG-biomarker literature, the precise model structure and weighting of individual predictors cannot be independently examined.

Finally, patients whose treatment was concordant with the report may have been managed by clinicians who are generally more engaged with biomarker-informed care, more systematic in monitoring, or able to invest more time in treatment planning, and these differences in care processes could contribute to the observed advantage of the concordant group.

These limitations highlight the need for prospective, adequately powered trials that randomize patients to biomarker-guided versus usual care conditions and standardize outcome assessments across treatment modalities. Such studies should also investigate potential moderators of benefit, including sex and gender, comorbidity profiles, baseline severity, prior treatment history and care setting, and should incorporate economic evaluations and patient-reported outcomes such as functioning, quality of life and treatment satisfaction to clarify real-world value. In addition, integrating clinical and questionnaire-based predictors with EEG/ECG features into unified decision models may further improve performance compared with neurophysiological data alone.

In its current form, the biomarker report reflects the presently available EEG/ECG prediction literature, but it should be regarded as a dynamic platform that can be iteratively refined as new biomarkers and treatment-specific predictors are validated. Over time, the incorporation of emerging evidence may allow more precise stratification and further enhance its value as a clinical decision-support tool.

Taken together, this observational real world analysis provides an initial indication that, within routine specialist care, treatments that align with an EEG/ECG based biomarker report are associated with higher response rates across several established interventions for depressive episodes. These findings are hypothesis generating and may support the rationale for more definitive prospective trials of biomarker guided, multimodal treatment strategies.

## Data Availability

All data produced in the present study are available upon reasonable request to the authors

## Conflicts of interest

M.d.B., S.O. and E.A. are cofounders and/or employees of DeepPsy AG. B.P. has received honoraria for lectures from SGIP-SSPI, Johnson & Johnson and Lundbeck. J.J. has received honoraria for lectures from Janssen Switzerland, SGIP-SSPI, Schwabe Pharma, Johnson & Johnson and Lundbeck. S.P. has received honoraria from Beltz and travel support from Schwabe Pharma. All other authors declare that they have no conflicts of interest.

## Funding source

CTI was supported by the University of Macau (File no: SRG2023-00040-ICI and MYRG-GRG2024-00022-ICI) and the Science and Technology Development Fund (FDCT) of Macau (0092/2025/ITP2).

## CRediT Author Statement

Barbora Provaznikova: investigation, writing - original draft, visualization. Mateo de Bardeci: methodology, validation, writing - review & editing, visualization. Enzo Altamiranda: data curation. Cheng-Teng Ip: methodology, review & editing. Anna Monn: validation, methodology, supervision, writing - review & editing. Samantha Weber: validation, writing - review & editing. Johannes Jungwirth: investigation, review & editing. Judith Rohde: writing - review & editing. Susanne Prinz: writing - review & editing. Golo Kronenberg: writing - review & editing. Annette Brühl: supervision, writing - review & editing. Tobias Bracht: supervision, writing - review & editing. Sebastian Olbrich: conceptualization, investigation, methodology, formal analysis, supervision, writing - review & editing.

## Acknowledgements

We would like to express our sincere gratitude to Ms. Eliane Strickler and her team for their invaluable support with the EEG recordings. We also thank the psychologists Ms. Yamina Ehrt-Schäfer, M.Sc., and Mr. Johannes Vetter, PhD, for their important contributions to the study and their help with recruitment. Our special thanks go to Ms. Angelina Frasch, administrative coordinator, and the entire secretary team for their highly appreciated organizational support.

## Notes

### Author Declarations

An ethical vote was obtained by the Swiss Association of Research Ethics Committees (Nr. 2023-00494), and ethical approval was granted (Nr. 2025-00559).

